# Mega-analytic support for Early Start Denver Model, age at intervention start, and pre-intervention developmental level as factors differentiating early intervention outcomes in autism

**DOI:** 10.1101/2025.04.14.25325786

**Authors:** Veronica Mandelli, Elena Maria Busuoli, Michel Godel, Nada Kojovic, Yana Sinai-Gavrilov, Tali Gev, AEIR Consortium, Annarita Contaldo, Eric Courchesne, Karen Pierce, Ofer Golan, Antonio Narzisi, Filippo Muratori, Costanza Colombi, Sally J. Rogers, Giacomo Vivanti, Marie Schaer, Liliana Ruta, Michael V. Lombardo

**Affiliations:** Laboratory for Autism and Neurodevelopmental Disorders, Center for Neuroscience and Cognitive Systems @UniTn, Istituto Italiano di Tecnologia, Rovereto, Italy; Department of Psychiatry, University of Geneva School of Medicine, Geneva, Switzerland; Department of Psychology, Bar-Ilan University, Ramat-Gan, Israel; IRCCS Stella Maris Foundation, Calambrone, Italy; Autism Center of Excellence, Department of Neurosciences, University of California, San Diego, La Jolla, CA, USA; MIND Institute, Department of Psychiatry, University of California, Davis, Davis CA, USA; A.J. Drexel Autism Institute, Drexel University, Philadelphia, PA, USA; Institute for Biomedical Research and Innovation (CNR-IRIB), National Research Council of Italy, Messina, Italy

**Author notes:** Corresponding author: Michael V. Lombardo. Equal first author contributions (Veronica Mandelli, Elena Maria Busuoli). The full list of AEIR Consortium authors can be found in the acknowledgments section. Equal senior author contributions (Annarita Contaldo, Eric Courchesne, Karen Pierce, Ofer Golan, Antonio Narzisi, Filippo Muratori, Costanza Colombi, Sally J. Rogers, Giacomo Vivanti, Marie Schaer, Liliana Ruta, Michael V. Lombardo).

## Abstract

**Objective:** Autism early intervention meta-analyses have provided initial answers to questions such as ‘*what types of interventions work*’ and ‘*for what outcomes*’? However, we also want to know ‘*for whom*’ is early intervention most effective for? Mega-analysis can offer up complementary insights to meta-analyses regarding the ‘*what works*’ and ‘*for what*’, while also offering unique insights into the ‘*for whom*’ question.

**Methods:** Here we conduct a mega-analysis with linear mixed effect modeling on AEIR consortium early intervention datasets totaling n=645 children spanning several countries (e.g., USA, Switzerland, Italy, Israel, and Australia). Early Start Denver Model (ESDM) and other non-ESDM approaches (e.g., EIBI, NDBI, other community/treatment as usual approaches) was evaluated as contrasting intervention types. Models also evaluated intervention intensity, type, participant sex, age at intervention start, and pre-intervention developmental quotient. Subscales of Mullen Scales of Early Learning (MSEL), Vineland Adaptive Behavior Scales (VABS), and Autism Diagnostic Observation Schedule (ADOS) were utilized as outcome measures.

**Results:** Neither intervention intensity nor participant sex affected outcomes. ESDM showed faster growth in language and non-verbal cognition compared to non-ESDM intervention. Irrespective of intervention type, earlier intervention start was associated with increased MSEL and VABS scores and decreased ADOS severity. Growth trajectories on the MSEL also varied by pre-intervention developmental quotient, with higher quotients predicting faster growth irrespective of intervention type.

**Conclusions:** Age at intervention start and pre-intervention developmental quotient are important individualized factors that predict early intervention response. ESDM also impacts language, non-verbal cognition, and core autism features.

---

Autism is one of the most common neurodevelopmental conditions in society today and represents a broad population of individuals that are heterogeneous across multiple scales, from genotype to phenotype (Lombardo et al., 2019), across development (Gentles et al., 2023), and across important clinical outcomes (e.g., responses to intervention) (Godel et al., 2022; Lombardo et al., 2021; Vivanti, Prior, et al., 2014). A top priority on the path towards precision medicine is the development of early intervention approaches that facilitate developmental gains and positive outcomes aligned with multiple different stakeholder perspectives (Lombardo & Mandelli, 2022; Pellicano & den Houting, 2021; Tager-Flusberg & Kasari, 2013). The push for earlier diagnosis and intervention is also paramount (Pierce et al., 2021; C. J. Smith et al., 2022) and predicated on the idea that there is higher probability of facilitating more positive outcomes because of the greater neural plasticity offered during the earliest periods of neurodevelopment (Dawson, 2008). The top priorities/questions for early intervention research are quite clear. We seek to better understand 1) *what works*, 2) *for whom*, and 3) *for what* (Sandbank, Bottema-Beutel, Crowley, Cassidy, Dunham, et al., 2020)? Since heterogeneity in response to early intervention is a high priority topic, it is important to further unpack the ‘*for whom*’ question. We need to better understand *how* and *why* early intervention is facilitating differential individualized outcomes. For these important questions, we have to go beyond examination of on-average group-differences due to intervention and dissect what are the factors or characteristics present in children before intervention begins that would help us predict the subsequent developmental path during a particular intervention (Lombardo et al., 2021; Mandelli et al., 2023; Vivanti, Prior, et al., 2014). Another important priority is to better understand what is changing in terms of underlying neurobiology as a function of the differential experience and learning provided by effective early intervention (Dawson et al., 2012; Jones et al., 2017; Lombardo et al., 2021).

Prior reviews and meta-analyses of autism early intervention research provide an initial starting point on the ‘*what works*’ and ‘*for what*’ questions. Such work helps us get a better sense of how early intervention may or may not have effects at a group-level. However, despite extensive culls through the literature on early intervention, results can seem mixed (e.g., (Crank et al., 2021; Eldevik et al., 2009; Fuller & Kaiser, 2020; Howlin et al., 2009; McGlade et al., 2023; Rodgers et al., 2021; Sandbank, Bottema-Beutel, Crowley, Cassidy, Dunham, et al., 2020; Sandbank, Bottema-Beutel, Crowley, Cassidy, Feldman, et al., 2020; Sandbank et al., 2023; Warren et al., 2011). For example, without filtering or controlling for study quality/bias, there is some evidence that various types of early intervention (e.g., developmental and naturalistic developmental behavioral interventions; NDBI) can be effective on-average in changing a variety of outcomes including language, intellectual and social communication abilities (Fuller et al., 2020; Sandbank, Bottema-Beutel, Crowley, Cassidy, Dunham, et al., 2020; Sandbank, Bottema-Beutel, Crowley, Cassidy, Feldman, et al., 2020; Sandbank et al., 2023). However, a somewhat different picture emerges when evaluating a smaller handful of high-quality randomized controlled trials (RCT). Using this particular restriction, earlier meta-analyses showed that several types of early behavioral interventions may be limited in effectiveness in changing cognitive, language, or core autism symptom domains (Crank et al., 2021; McGlade et al., 2023; Sandbank, Bottema-Beutel, Crowley, Cassidy, Dunham, et al., 2020). However, a more recent updated meta-analysis suggests that some intervention types like naturalistic developmental behavioral interventions (NDBI) (e.g., Early Start Denver Model, ESDM) can promote positive outcomes in cognitive, language, adaptive functioning, and core diagnostic characteristics of autism (Sandbank et al., 2023).

While meta-analytic inferences are very important, there may also be some limitations for some research priorities/questions. First, meta-analytic work enables tests of whether replicable non-zero group-level effects exist in the literature. This goal is tailored to answer the ‘*what works*’ and ‘*for what*’ questions quite well. Nevertheless, these goals may be complicated when heterogeneity in response to intervention exists (i.e. answering the ‘*for whom*’ question). Although evidence from high-quality RCTs suggests that group-level effects vary considerably across the literature (Crank et al., 2021; McGlade et al., 2023; Sandbank, Bottema-Beutel, Crowley, Cassidy, Dunham, et al., 2020; Sandbank et al., 2023), this does not preclude the fact that the interventions may still have important effects on specific types of individuals. Thus, a different analytic approach may be complementary for answering the ‘*for whom*’ question. Approaches such as individual participant data meta-analysis (IPD-MA) or ‘mega-analysis’ (Eisenhauer, 2021) may be complementary to meta-analysis by allowing for deeper insight than what may be possible from meta-analysis alone. Mega-analysis utilizes the raw data from individual participants across many studies and allows for testing not only group-level effects, but also factors that explain individual differences in intervention response. To our knowledge, the only IPD-MA/mega-analysis in the literature (Rodgers et al., 2021) compares early intensive applied behavior analysis (ABA) based interventions, such as variants of early intensive behavioral intervention (EIBI) and NDBI, to treatment-as-usual (TAU) or eclectic interventions. The primary findings here suggest that early intensive ABA-based interventions have a small effect on intellectual and adaptive functioning. Other moderating individual difference factors such as sex, age, baseline cognitive or adaptive functioning level, did not interact with intervention type to explain heterogeneity in intervention response.

A second limitation of meta-analytic work is that statistical power to detect on-average group-level effects may be low when restrictions are placed on filtering for only a small subset of high-quality studies. Indeed, in contrast to earlier meta-analyses (Sandbank, Bottema-Beutel, Crowley, Cassidy, Dunham, et al., 2020), the more recent updated meta-analyses with higher statistical power via the inclusion of more studies, tends to reveal somewhat difference inferences (Sandbank et al., 2023). Additionally, the small handful of existing high-quality RCT studies tend to have relatively small sample sizes. In the case of small sample sizes, unadjusted descriptive statistics and standardized effects size computed from them will be inherently less precise and possibly more prone to effect size inflation (Lombardo et al., 2019). Utilizing unadjusted statistics in meta-analysis is a necessary step to homogenize effect size computation across heterogeneous studies. However, this attribute does not allow for correction for covariates within-study that would typically be applied in each individual study. To tackle this issue to some extent, moderator analyses from meta-regression models can be implemented to test associations with study-specific factors and covariates (e.g., mean age at intervention start). These moderator analyses are limited though to study-specific summary factors (e.g., mean age) that themselves are descriptive statistics computed per study and are inherently not the same measuring and controlling for those covariates within individuals (e.g., age at intervention start per individual). Again, utilization of mega-analysis addresses this limitation by allowing for covariates to be applied within-study at the level of individuals.

To overcome these limitations, we developed the Autism Early Intervention Research (AEIR) consortium to pool together individual participant data from many studies on early intervention in autism for the purpose of conducting mega-analyses. AEIR has been able to pool together 645 autistic individuals with age range between 13 to 60 months that can be reduced to two main intervention types: Early Start Denver Model (ESDM) and non-ESDM high quality early interventions approaches (e.g., ABA, PRT, STAR) as well as interventions employed in community settings and commonly referred ‘treatment-as-usual’ (e.g., speech, occupational, psychomotor therapy). A contrast of ESDM versus non-ESDM is utilized in this work for sake of convenience given the roughly half split between these two intervention approaches within the AEIR consortium dataset (Figure 1; Supplementary Table 1). Contrasting ESDM to non-ESDM approaches in a mega-analysis can be useful for comparing to other meta-analyses (Fuller et al., 2020) as well as previous individual high-quality ESDM RCT studies (Dawson et al., 2010; Estes et al., 2015; Rogers et al., 2019, 2021). In addition to contrasting the effects of ESDM vs non-ESDM intervention types, we also evaluate main effects and interactions between factors such as age, intervention intensity, age at intervention start, and pre-intervention developmental quotient as possible factors that can predict significant variance in intervention responses.

**Figure 1:**
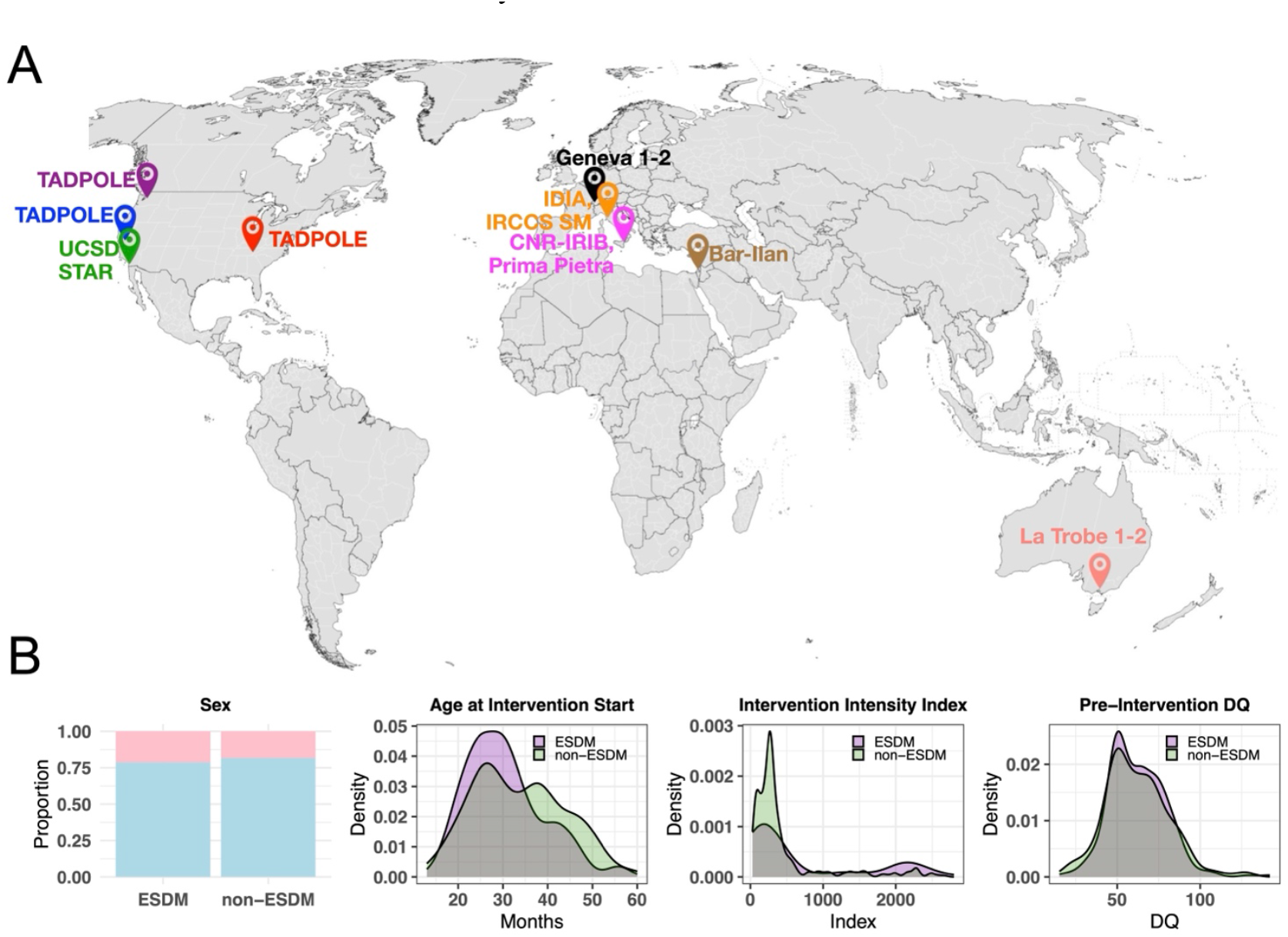
The Autism Early Intervention Research (AEIR) consortium along with intervention and characteristics describing the combined AEIR dataset. Panel A shows sites around the world where data from the AEIR consortium originates from. On the far left of Panel B is a plot that shows the proportion of males (blue) versus females (pink) broken down by the two intervention types (ESDM, left; non-ESDM, right). The plot on the mid-left shows age at intervention start density plots for the two intervention types (ESDM, purple; non-ESDM, green), where age at intervention start is indicated in months on the x-axis. The plot on the mid-right shows the intervention intensity index (see Methods for a description of how this index is computed) for the two intervention types. The plot on the far right shows pre-intervention developmental quotients (DQ; x-axis) for the two intervention types.

## Methods

All secondary data analysis reported here was approved by the Province of Trento Azienda Provinciale per i Servizi Sanitaria (APSS) ethical committee.

## Datasets

Data utilized in this study come from 11 international datasets on early intervention in autistic toddlers as part of the Autism Early Intervention Research (AEIR) consortium. Many, though not all, of these datasets have already been previously published independently (Bacon et al., 2014; Colombi et al., 2018; Contaldo et al., 2020; Godel et al., 2022; Muratori et al., 2014; Robain et al., 2020; Rogers et al., 2019; Sinai-Gavrilov et al., 2020; Vivanti et al., 2019; Vivanti, Paynter, et al., 2014). The final dataset comprises n=645 (female n=128) autistic children, with ages at intervention start ranging from between 13-60 months. All datasets have at least 2 timepoints within an individual but some datasets possess more than 2 timepoints per individual. See Supplementary Table 1 for characteristics of individual datasets (Fig 1A).

## Early Intervention Programs

Participants received between 3-27 months of early intervention from varying approaches and variable levels of intervention intensity (Supplementary Table 1). Given the considerable number of participants that completed one type of early intervention known as Early Start Denver Model (ESDM) we partitioned the data into two groups: 1) ESDM (n=304, females= 65) or 2) non-ESDM intervention (non-ESDM; n=341, females= 63). The non-ESDM interventions represents a combination of other types of early interventions commonly used in the literature and in community settings (e.g., speech therapy, occupational therapy, ABA/Discrete Trial Training, Pivotal Response Training, STAR program, etc). Early interventions varied in their format of administration and could be comprised of individual, group, and parent intervention components.

## Outcome measures

All datasets had commonalities with reference to measures utilized to examine cognitive ability, language, motor, adaptive functioning, and autism symptom severity. For cognitive ability, language, and motor skills, most datasets in AEIR, with the exception of those originating in Italy, utilized the Mullen Early Scales of Learning (MSEL) (Mullen, 1995). Scores on the MSEL that were utilized were subscales of expressive and receptive language (EL, RL), visual reception skills (VR) and fine motor (FM). For these subscales, we utilized age-equivalent scores in our developmental trajectory analyses to examine how outcome measures change over age and scale with changes in age-equivalent score growth. Adaptive functioning was measured in all datasets with the Vineland Adaptive Behavior Scales (VABS) (Sparrow et al., 2005). For the VABS we utilized standardized scores on domains such as Communication, Socialization, Daily Living Skills, and Motor. Finally, concerning autism symptom severity, here we utilized the Autism Diagnostic Observation Schedule (ADOS) Calibrated Severity Scores (CSS) (Esler et al., 2015; Gotham et al., 2009; Hus et al., 2014). ADOS CSS scores are advantageous because they are standardized relative to a child’s age and language ability and ensure comparability across ADOS administration modules.

## Predictor Measures

In this work we conducted a mega-analysis to clarify the main effects and interactions between intervention and subject characteristics, aiming to disentangle the contribution of each aspect. To achieve this, we needed to extract measures that were common across all datasets to use as predictors in our analysis. Regarding intervention-specific factors potentially able to predict intervention outcome measures, we assessed type of early intervention (ESDM or non-ESDM) and an intervention intensity index. The intervention intensity index we use helps to balance intensity formalized as hours and weeks of intervention with the number of adults and children present in the intervention context, and is computed as: (weeks of intervention * hours per week * number of adults) / number of children present (Waizbard-Bartov et al., 2021). Concerning the child characteristics at intervention start, we analyzed cognitive abilities at intervention start reported as developmental quotient scores (DQ). DQ scores were derived from the MSEL early learning composite (ELC) score. The exception here was for Italian datasets whereby MSEL was not collected. In this situation, DQ scores were derived from the Griffith Mental Development Scales (GMDS) (Griffith et al., 2006). Finally, age at intervention start and biological sex were also selected and analyzed as demographic factors that might be predictors associated with intervention responses.

## Statistical Analyses

In the first set of analyses, we evaluated whether the initial pre-intervention characteristics of children or intervention intensity were comparable between the ESDM and non-ESDM groups. To examine this, we employed a linear mixed effect model with fixed effects of intervention type, sex, and their interaction and with dataset modeled as random effect. Additionally, we utilized a chi-squared test to investigate whether the distribution of males and females varied by intervention type (Fig 1B).

In the second set of analyses, we aimed to test the influence of the predictors on intervention outcomes. We utilized a set of linear mixed effect models to account for repeated measures and handle nested within-study factors. Three sets of models were analyzed, organized by whether the outcome measures were from MSEL, VABS, or ADOS. For MSEL models, the dependent variable was the age-equivalent scores of each of the MSEL subscales, with one model run for each subscale. Datasets from Italy were excluded from these models as these datasets did not have MSEL data available. For VABS models, the dependent variable was the standardized domain scores, with a separate model for each VABS domain. For the ADOS we employed three distinct models, with each model analyzing a different CSS score as the dependent variable. In terms of the independent variables, we used the same variables for all models to enable comparison. Fixed effects in the models were age, intervention type, intervention intensity index, sex, age at intervention start, and pre-intervention developmental quotient. Interaction effects were also modeled as fixed effects and include interactions between age and pre-intervention DQ, age at intervention start, and intervention type. Random effects were also utilized in the model to account for subject-specific age intercepts and slopes (i.e. age modeled within subject-ID, dataset). All linear mixed effect modeling was conducted in R using the *lmer* function within the *lmerTest* R library. To identify significant effects across multiple analyses, p-values were utilized to compute FDR values, and then FDR control for multiple comparisons was achieved with thresholding at q<0.05. Code implementing all analyses can be found at https://gitlab.iit.it/bmp006-public/LAND_IIT/aeir1.

## Results

### Pre-intervention differences between ESDM and non-ESDM

In this mega-analysis, we report data compiled from the AEIR consortium across 11 datasets (Figure 1A) comprising n=645 autistic individuals. Data were split into two main types of early intervention: 1) ESDM (n=304, male = 78.6%, age range = 14-57 months) and 2) non-ESDM (n=341, male = 81.5%, age range = 13-60 months). See Supplementary Table 2 for a breakdown of descriptive statistics of baseline measures between these intervention types. Intervention types did not differ in proportion of males versus females (*χ*^*2*^ = 0.68, *p* = 0.40). After controlling for dataset, pre-intervention developmental quotient did not differ between intervention types (*F(1,569)* = 2,6, *p* = 0.23). Age at intervention start differed between intervention types (*F(1,637)* = 32.7, *p* = 4.72e-08), with ESDM showing on average younger ages than non-ESDM by 3 months (mean age, ESDM = 30.46 months; non-ESDM = 33.61). Intervention intensity index also differed between intervention types (*F(1,637)* = 66.8, *p* = 4.66e-15) with ESDM being on-average more intense than non-ESDM (Supplementary Table 3). Given these differences between intervention types, these variables are included in all further models to account for variability attributed to them.

### Effects of intervention type

A primary research priority for early intervention is to examine questions of ‘*what works*’ and ‘*for what*’. Here we assess those questions specifically with a contrast of ESDM versus non-ESDM intervention types. If intervention type differs, we would expect an intervention type*age interaction effect, whereby group-level trajectories of one intervention type significantly differ compared to the other intervention type (Fig 2A). Using ADOS CSS scores as the outcome measure, we find an intervention type*age interaction specifically for ADOS RRB CSS (*F* = 5.71, *p* = 0.0171, *FDR q* = 0.0385) whereby ESDM has a slightly steeper trajectory (i.e. increasing severity over time) than non-ESDM (Fig 2B). Such interactions with age were not found for ADOS CSS SA or total CSS scores. Using the VABS as the outcome measure, we did not find any intervention type*age interactions for any of the VABS domains (Fig 2C). Thus, ESDM and non-ESDM largely have relatively similar rates of growth over development for the VABS. Using the MSEL as the outcome measure, we detected a significant intervention type*age interaction for receptive language (RL) (*F* = 8.28, *p* = 4.16e-3, *FDR q* = 7.49e-3) (Fig 2D). This effect can be described as ESDM showing a slightly steeper trajectory (i.e. faster growth) compared to non-ESDM. Similar effects of faster growth for ESDM compared to non-ESDM are evident for expressive language (EL) and visual reception (VR) subscales, when a more liberal false discovery rate (FDR) threshold of q<0.1 is applied to the intervention type*age interaction (EL: *F* = 4.19, *p* = 0.0412, *FDR q* = 0.0742; VR: *F* = 5.27, *p* = 0.0395, *FDR q* = 0.0711). In contrast, no intervention type*age interaction is present for the MSEL FM subscale. See Supplementary Table 4 for the full set of statistics for all models and comparisons in Figure 2.

**Figure 2:**
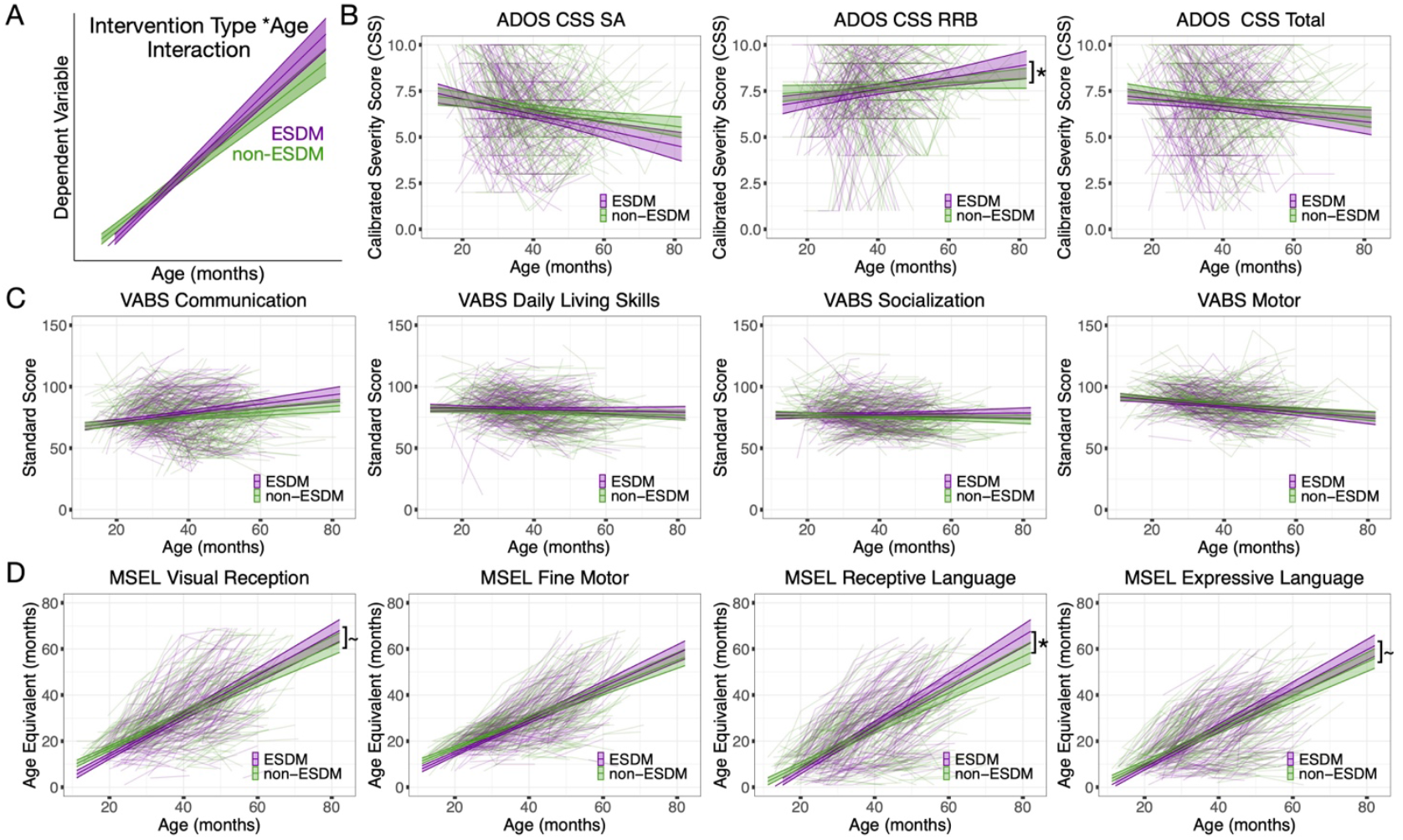
Comparison of ESDM versus non-ESDM intervention. Panel A is a hypothetical figure showing an example of a significant intervention type*age interaction effect, whereby one intervention type (e.g., ESDM) shows faster growth over time than the other intervention type (e.g., non-ESDM). Panels B-D show spaghetti plots for ADOS (B), VABS (C) and MSEL (D) outcome measures. In all plots, ESDM is shown in purple, while non-ESDM is shown in green. The x-axis shows chronological age in months, while the y-axis shows the dependent variable for each measure (e.g., CSS scores for ADOS, standardized scores for VABS, age-equivalent scores for MSEL). Each individual child is shown with transparent lines, while the group-level trajectory along with 95% confidence bands are shown overlaid on top. The ∼ symbol indicates intervention type*age interaction effects passing FDR q<0.1, while a star (*) indicates intervention type*age interaction effects that passing FDR q<0.05.

### Effects of age at intervention start

While answering the ‘*what works*’ and ‘*for what*’ is fundamentally important, there is also the important question of ‘*for whom and how*’ does early intervention work best for individuals? To answer these questions, here we report results from our models for predictor variables such as sex, age at intervention start, and pre-intervention developmental quotient. Sex has no effect for nearly all MSEL, VABS, or ADOS dependent variables, apart from a small effect (i.e. Male>Female) for VABS motor (*F* = 4.99, *p* = 0.026, *FDR q* = 0.039). In contrast, there is a main effect for age at intervention start, which can be described as generally having better scores most outcome measures (i.e. higher VABS and MSEL scores, lower ADOS scores) for individuals that start intervention earlier (Fig 3A). This is clearly shown in Figure 3B for the VABS and MSEL, whereby individuals starting intervention earlier (e.g., depicted as progressively lighter blue across deciles defined by age at intervention start) have on-average higher scores both at entry and on the outcome measure. These effects are most evident on MSEL and VABS, as these main effects of age at intervention start are evident without an interaction between age at intervention start and age (Supplementary Table 4). An exception to this statement occurs for the VABS Motor subscale, because there is a significant age*age at intervention start interaction effect (F = 7.85, p = 0.00542, FDR q = 0.0122) described by declining motor trajectories in individuals that start earlier followed by later flattening of trajectories in individuals starting intervention later (Fig 3B). Age*age at intervention start interaction effects also occur for ADOS SA CSS scores (F = 8.03, p = 0.00472, FDR q = 0.0106) and to a lesser extent for ADOS CSS total scores (F = 4.51, p = 0.0338, FDR q = 0.06). These interactions are described by declining trajectories in individuals starting earlier (i.e. decreasing autism severity over time), while trajectories flatten out and become more stable over time for individuals who start intervention later (Fig 4A). See Supplementary Table 4 for all statistics for these effects across all models.

**Figure 3:**
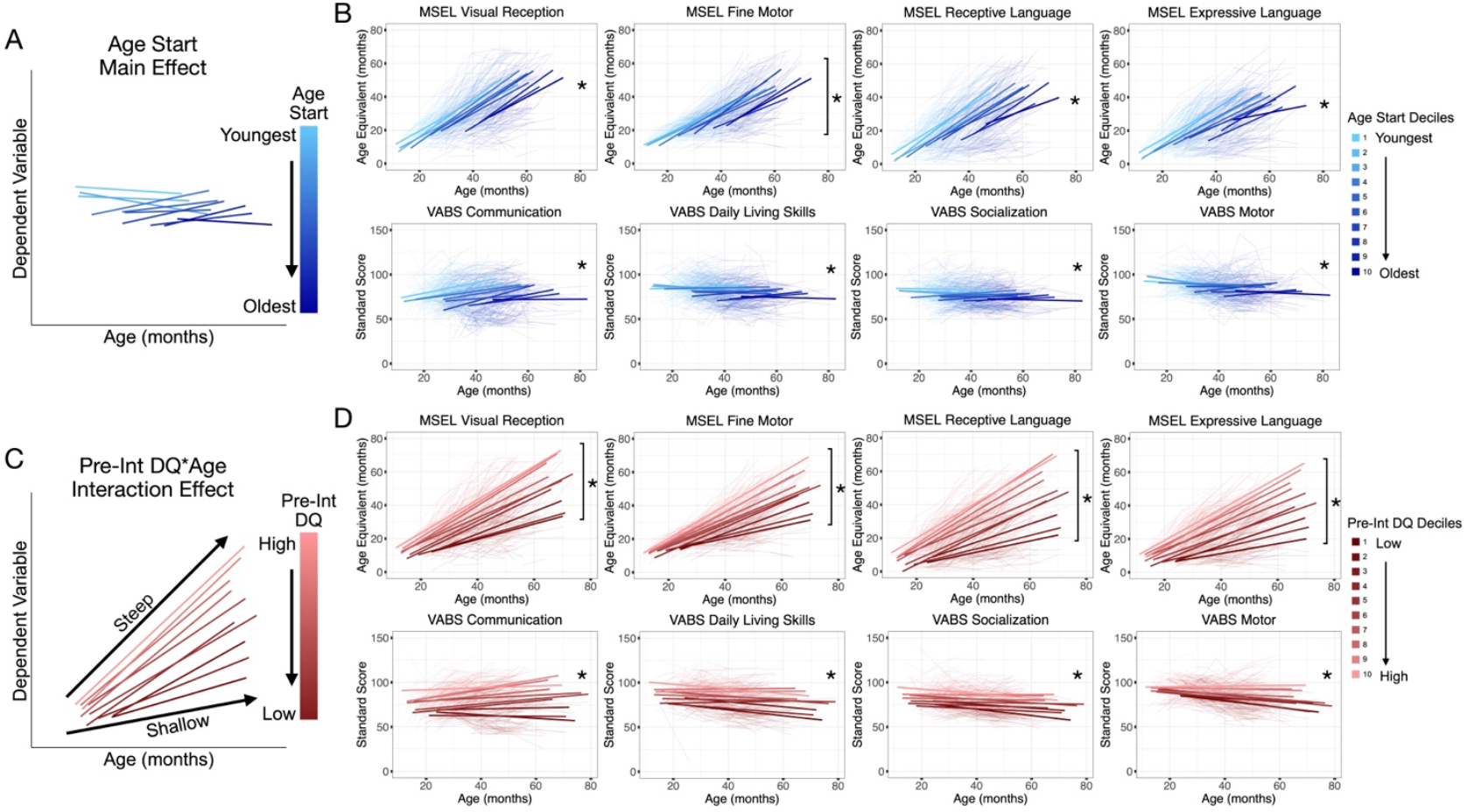
Effect of age at intervention start and pre-intervention developmental quotient on Mullen (MSEL) and Vineland (VABS) scores. Panel A and C show hypothesized examples of a main effect of age at intervention start (A) and an interaction between pre-intervention developmental quotient (Pre-Int DQ) and age (C). The coloring in all plots indicates continuous variation in each decile of the main predictor variable. For age at intervention start, the lowest deciles (lighter blue) are for individuals starting intervention at earlier ages. Increasing deciles (progressively darker blue) scale with progressively later age at intervention start. For Pre-Int DQ, the lowest deciles (darker red) are for individuals with the lowest Pre-Int DQ. Increasing deciles (progressively lighter red) scale with increasing levels of Pre-Int DQ. The plot in panel A shows a hypothetical main effect of age at intervention start, whereby there generally higher scores are observed for individuals that start intervention at the earliest ages. Note that there is no interaction with age, and this indicates a situation where the trajectories stay parallel as one increases from starting earlier to later. In contrast, Panel C shows a hypothetical interaction between Pre-Int DQ and age. In this situation, the slope of trajectories change as one goes from lower Pre-Int DQ (e.g., shallower slopes indicative of relatively slower growth) to higher Pre-Int DQ (e.g., steeper slopes indicative of relatively faster growth). Panels B and D show spaghetti plots of the actual data. Chronological age in months is plotted on the x-axis, while the outcome dependent variable is plotted on the y-axis (e.g., standardized scores for VABS, age-equivalent scores for MSEL). Individuals are shown with the transparent lines and decile trajectories are shown with thicker lines overlaid on top. Panels B and D shows the effects of age at intervention start (B) and Pre-Int DQ (D) on Mullen (MSEL, top) or Vineland (VABS, bottom). In panel B all outcome measures show a significant main effect of age at intervention start (e.g., indicated with the * for a significant effect passing FDR q<0.05), indicative of a stacking pattern with lighter blue deciles of individuals that started treatment earlier tending to have generally higher scores on the outcome measures. The Mullen Fine Motor also has a significant age at intervention start by age interaction (e.g., indicated by the star and brackets,]*, passing FDR q<0.05). This effect can be explained by flatter slopes for individuals starting intervention earlier and steeper slopes for individuals that started later. Panel D shows Pre-Int DQ interaction with age for MSEL outcomes (e.g.,]*) and a main effect of Pre-Int DQ with no age interaction for VABS outcomes.

**Figure 4:**
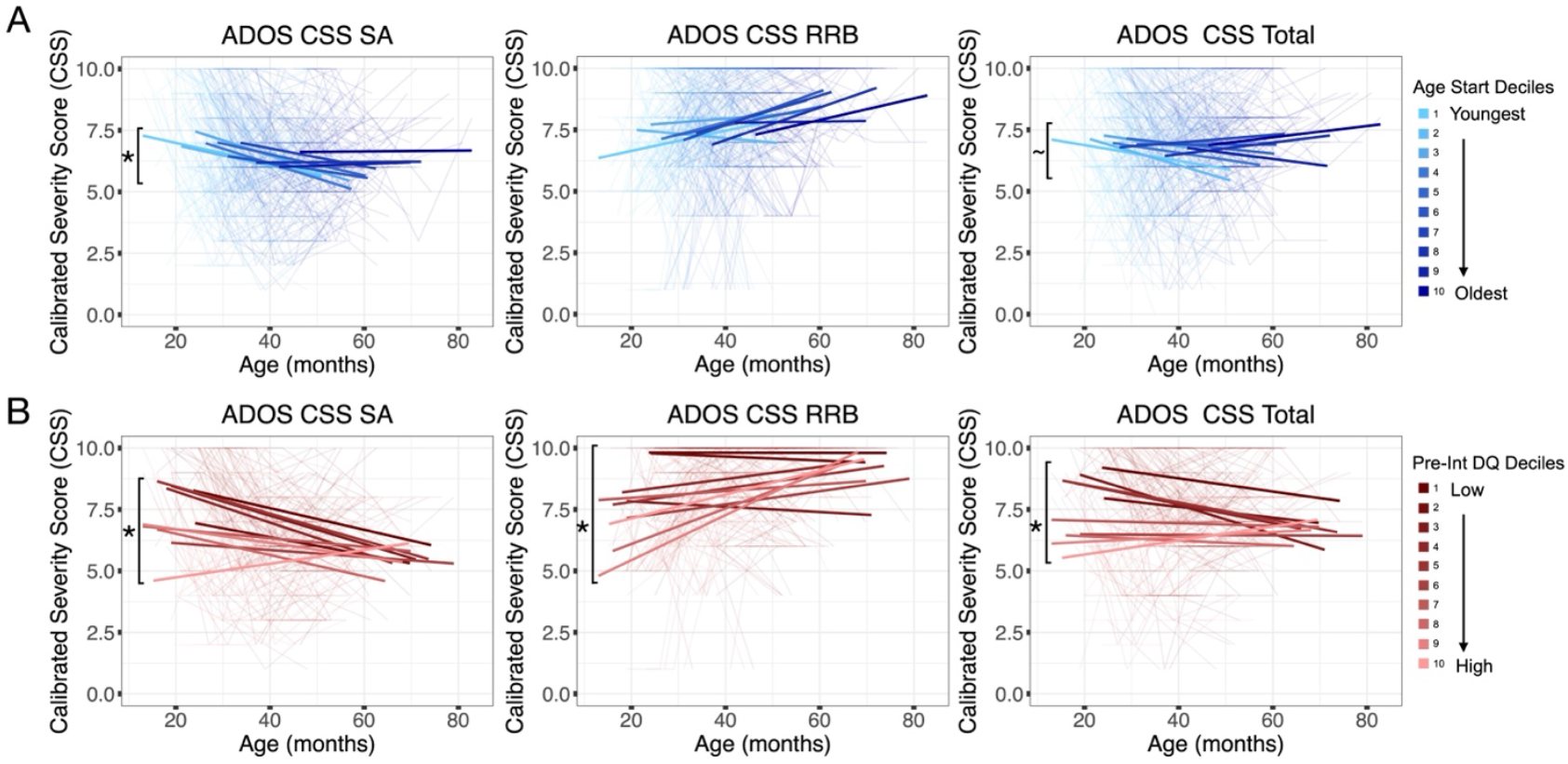
Effects of age at intervention start and pre-intervention developmental quotient on autism calibrated symptom severity (ADOS CSS) scores. The plots in this figure depict the effects of age at intervention start (A) or pre-intervention developmental quotient (Pre-Int DQ) (B) on ADOS CSS SA (left), CSS RRB (middle) and ADOS CSS Total scores (right). In these spaghetti plots, the x-axis shows chronological age in months, while the y-axis shows ADOS CSS scores from 0-10, with 0 being least severe and 10 being most severe. Individuals are shown with the transparent lines, while decile trajectories are shown with thicker lines overlaid on top. Interaction effects between the predictor variable and age are annotated with a star and bracket (e.g.,]*) or ∼ and bracket (e.g.,]∼). A tilde symbol (e.g., ∼) represents an effect passing FDR q<0.1, while the star (*) represents a significant effect passing FDR q<0.05. The coloring in all plots indicates continuous variation across deciles of the predictor variable. For age at intervention start, the lowest deciles (lighter blue) are for individuals starting earliest, with increasing deciles (darker blue) scaling with later age at intervention start. For Pre-Int DQ, the lowest deciles (darker red) are for individuals with the lowest Pre-Int DQ and increasing deciles (lighter red) scale with increasing levels of Pre-Int DQ.

### Effects of pre-intervention developmental quotient

While intervention type and age at intervention start exert important effects on intervention responses, perhaps the strongest predictors of all are the effect of pre-intervention developmental quotient (Pre-Int DQ). Main effects of Pre-Int DQ are ubiquitous across all MSEL, VABS, and ADOS outcome measures. Individuals with generally higher Pre-Int DQ tend to have on-average higher scores on the developmental outcome measures (Supplementary Table 4). On the MSEL and ADOS (but not VABS) there is also evidence of an interaction between Pre-Int DQ and age, indicating differential developmental trajectories that vary as a function of Pre-Int DQ. This interaction effect for the MSEL is described as a ‘fanning out’ effect – individuals with the highest Pre-Int DQ show greater increases in developmental age over time than individuals with the lowest Pre-Int DQs, though those individuals also demonstrate an accelerating learning slope over time (Fig 3C-D). For ADOS SA and Total CSS scores, the interaction between Pre-Int DQ and age for individuals with the lowest pre-intervention developmental quotients start with very high severity CSS scores, and over time demonstrate decreasing severity of CSS scores. In contrast, those with higher pre-intervention developmental quotient individuals demonstrate little change over time in mid-severity CSS scores, which are similar at the endpoint to those of the individuals with lower pre-intervention developmental quotients (Fig 4B). Such effects can be interpreted differently than the RRB interaction effects, whereby flat trajectories in low Pre-Int DQ individuals but increasing in high Pre-Int DQ individuals. See Supplementary Table 4 for all statistics for these effects across all models.

## Discussion

In this work we present insights from a mega-analysis of autism early intervention datasets from the AEIR consortium. This mega-analysis was positioned to facilitate answers to primary questions within research on early intervention in autism - that is, *what works, for whom*, and *for what* (Sandbank, Bottema-Beutel, Crowley, Cassidy, Dunham, et al., 2020). To target *what works* and *for what* questions, we focused on the contrast of one of ESDM, as one promising naturalistic developmental behavioral intervention (NDBI), versus other types of EIBI and NDBI early interventions or other community-based treatment-as-usual options. Congruent with some meta-analytic inference on ESDM (Fuller et al., 2020), we find there is some evidence that ESDM has beneficial effects on development of skills, particularly in receptive language (MSEL RL). These benefits manifest after controlling for a variety of other factors in the model, such as variability inherent to different datasets, intervention intensity, sex, age at intervention start, and pre-intervention developmental quotient. While our observations are not couched solely within large and well-controlled randomized double-blind studies, they complement insights from meta-analyses (Fuller et al., 2020; Sandbank, Bottema-Beutel, Crowley, Cassidy, Dunham, et al., 2020; Sandbank et al., 2023) that also describe some beneficial effects for NDBIs like ESDM when compared to other types of community early interventions. Despite this, the spaghetti plots shown in Figure 2 reveal considerable variability at the level of individuals that is not easily explained by intervention type. This observation underscores the fact that while ESDM may be on-average promoting some developmental skills more effectively than other non-ESDM intervention approaches, answers to the ‘*for whom*’ question will likely be important for closing the gap relating to this unexplained variance (e.g., (Godel et al., 2022; Lombardo et al., 2021)).

One of the key insights enabled by our mega-analysis relative to meta-analysis or prior smaller scale individual studies is the ability to isolate individualize factors and their key role in moderating developmental trajectories throughout the course of early intervention. Several factors tested here in this mega-analysis have been previously theorized as being potentially important individualized moderators of intervention response, particularly age at intervention start, pre-intervention developmental quotient, intervention intensity, and sex (Ben-Itzchak et al., 2014; Dawson, 2008; Godel et al., 2022; Rogers et al., 2021; Sandbank et al., 2024; T. Smith et al., 2015; Vivanti, Prior, et al., 2014). A characteristic separating this work from past work is that most past studies were individually of small sample size. Thus, this work benefits from pooling across datasets and increases statistical power and can make for a more definitive statement about the presence or absence of such effects on early intervention response. Here we find ubiquitous and strong impact for two variables: age at intervention start and pre-intervention developmental quotient across all variables examined (MSEL, VABS, ADOS). For age at intervention start, the main effect of this variable can be interpreted as higher scores on outcomes for individuals that started intervention earlier. Age at intervention start also interacted with age for the ADOS, with individuals who started intervention earlier showing some significant declining trajectories in symptom severity compared to individuals who started intervention relatively later. This effect could be congruent with some meta-analytic inference suggesting that NDBI interventions have a small effect on autism symptomatology (Sandbank et al., 2023). Qualifying this interpretation, our study shows that such an effect may be most present in individuals that start intervention at earlier timepoints. These observations are highly relevant to the discussion centered around the need for earlier diagnosis in order to begin interventions earlier (Dawson, 2008; Lombardo et al., 2021; Pierce et al., 2021; Zwaigenbaum et al., 2015). The theoretical idea behind this push is to capitalize on greater earlier neural plasticity to facilitate developmental gains for children (Dawson et al., 2012; Jones et al., 2017). Our findings illustrating the important impact age at intervention start has on intervention outcomes and is consistent with this idea. Overall, our results provide strong and definitive support for advocacy for earlier diagnosis and intervention, as age at intervention start was one of the most important moderating factors we found that positively influenced an individual’s response to intervention.

For pre-intervention developmental quotient, we found that this individualized characteristic interacted with age, indicating differential developmental trajectories depending on the child’s general level of ability pre-intervention. While all children are growing on MSEL subscales, the rates of growth tend to be steeper for children that have higher pre-intervention developmental quotients (Figure 3D). It is notable here that MSEL subscales measure important developmental features in children that are generally considered as features that are outside of the core of autism (e.g., language, motor, intellectual functioning). When considering core autism features, as measured by the ADOS, we find effects that are fundamentally different than those observed on the MSEL. Here, we find that individuals with the lowest pre-intervention developmental quotients tended to show improving ADOS severity levels and these trajectories differed from the flat to slightly increasing severity over time exhibited by individuals with higher pre-intervention DQ (Figure 4B). This effect on the ADOS suggests that interventions can still have considerable impact on core autism features, particularly for individuals that start intervention at lower levels. This type of effect on ADOS SA and Total CSS scores, but not the RRB is also important. Early interventions tend to target SA abilities more than early RRB, and thus the potentially important change that may facilitate improvements may be specific to SA behaviors targeted by early intervention. Overall, these results show promise for utilizing individualized characteristics to predict intervention outcomes and illustrates how core versus non-core features in autism may be differentially affected in individuals as a function of pre-intervention developmental level.

There are several caveats and limitations to underscore. First, the current mega-analysis was not restrictive in its inclusion criteria and as such, many of the contributing datasets may not meet the high standards of being well-controlled randomized double-blind studies. While such studies are a gold standard for evaluating interventions, the broader inclusion of other studies allowed for much wider reach and potentially enhanced generalizability to what is the reality in most community settings. The inferences observed in the current work on the effect of intervention type are however, consistent with most recent meta-analytic inferences about potential beneficial effects of NDBIs like ESDM (Fuller et al., 2020; Sandbank et al., 2023). Because the inclusion of datasets did not attempt to a priori equate certain variables, we found that intervention type significantly differed on variables such as age at start and intervention intensity. Although our statistical approach accounted for variance due to such variables in the model, further work contrasting intervention types with similar a priori control over age at start and intervention intensity would be needed to bolster inferences from such results. Second, our contrast tailored to assess the ‘*what works*’ question was limited to an assessment of how ESDM compares to a range of other diverse types of non-ESDM interventions that range from high-quality EIBIs and NDBIs to other approaches commonly used in the community and which target aspects that are not necessarily the core features of autism (e.g., speech, occupational, psychomotor therapy). Not enough datasets exist within the AEIR consortium for a full comparison of individual intervention types within the non-ESDM grouping we used for the current analysis. Thus, while the results suggest some facilitative effects of ESDM over non-ESDM interventions, this does not imply that other non-ESDM interventions were not also having a positive impact on a child’s progress. Third, although we have tested for effects like sex, due to the still limited number of actual females (e.g., around 60 individuals per each intervention type), statistical power to robustly test for sex differences in this study may be limited. Fourth, although we did not find an effect for intervention intensity, this null effect must be taken with caveat that intensity was not a variable that could be systematically manipulated and evaluated. Future work with more careful manipulations and variability in intervention intensity would be necessary to fully test whether this variable has important impact on outcomes.

In summary, using mega-analysis we have highlighted the prominent impact of two key factors: age at intervention start and pre-intervention developmental quotient, as moderators and predictors of individual differences in children’s response to early intervention. Second, we contrasted outcomes based on two different intervention types and found evidence for positive effects on receptive language, expressive language, and visual reception for ESDM compared to effects of non-ESDM interventions. Third, we found little evidence for effects related to a child’s sex or intervention intensity. These null findings must be taken with the caveat that both variables may not have been ideal (e.g., low power for females, lack of systematically varying intensity levels) in the current dataset. Finally, our work provides definitive strength and support to the knowledge base underlying some key factors that explain heterogeneity in intervention response.

## Data Availability

All data produced in the present study are available upon reasonable request to the authors

## Acknowledgments

We thank all participants and their families for participating in this study. The members of the Autism Early Intervention Research (AEIR) Consortium are (in alphabetical order): Elena Maria Busuoli, Costanza Colombi, Annarita Contaldo, Eric Courchesne, Tali Gev, Michel Godel, Ofer Golan, Nada Kojovic, Michael V. Lombardo, Veronica Mandelli, Filippo Muratori, Antonio Narsizi, Karen Pierce, Sally J. Rogers, Liliana Ruta, Marie Schaer, Yana Sinai-Gavrilov, Giacomo Vivanti.

## Author Contributions

Conceptualization: MVL. Methodology: MVL, VM, EMB. Formal analysis: MVL, VM, EMB. Investigation: MVL, VM, EMB, MG, NK, YSG, TG, AN, AC, CC, FM, OG, EC, KP, SJR, GV, MS, LR, AEIR. Writing - original draft preparation: MVL, VM, EMB. Writing - review and editing: MVL, VM, EMB. Visualization: MVL, VM. Supervision: MVL. Project administration: MVL. Funding acquisition: MVL.

## References

Bacon, E. C., Dufek, S., Schreibman, L., Stahmer, A. C., Pierce, K., & Courchesne, E. (2014). Measuring Outcome in an Early Intervention Program for Toddlers with Autism Spectrum Disorder: Use of a Curriculum-Based Assessment. Autism Research and Treatment, 2014, 1–9. 10.1155/2014/964704

Ben-Itzchak, E., Watson, L. R., & Zachor, D. A. (2014). Cognitive ability is associated with different outcome trajectories in autism spectrum disorders. Journal of Autism and Developmental Disorders, 44(9), 2221–2229. 10.1007/s10803-014-2091-0

Colombi, C., Narzisi, A., Ruta, L., Cigala, V., Gagliano, A., Pioggia, G., Siracusano, R., Rogers, S. J., Muratori, F., & Prima Pietra Team. (2018). Implementation of the Early Start Denver Model in an Italian community. Autism: The International Journal of Research and Practice, 22(2), 126–133. 10.1177/1362361316665792

Contaldo, A., Colombi, C., Pierotti, C., Masoni, P., & Muratori, F. (2020). Outcomes and moderators of Early Start Denver Model intervention in young children with autism spectrum disorder delivered in a mixed individual and group setting. Autism: The International Journal of Research and Practice, 24(3), 718–729. 10.1177/1362361319888344

Crank, J. E., Sandbank, M., Dunham, K., Crowley, S., Bottema-Beutel, K., Feldman, J., & Woynaroski, T. G. (2021). Understanding the Effects of Naturalistic Developmental Behavioral Interventions: A Project AIM Meta-analysis. Autism Research: Official Journal of the International Society for Autism Research, 14(4), 817–834. 10.1002/aur.2471

Dawson, G. (2008). Early behavioral intervention, brain plasticity, and the prevention of autism spectrum disorder. Development and Psychopathology, 20(3), 775–803. 10.1017/S0954579408000370

Dawson, G., Jones, E. J. H., Merkle, K., Venema, K., Lowy, R., Faja, S., Kamara, D., Murias, M., Greenson, J., Winter, J., Smith, M., Rogers, S. J., & Webb, S. J. (2012). Early behavioral intervention is associated with normalized brain activity in young children with autism. Journal of the American Academy of Child and Adolescent Psychiatry, 51(11), 1150–1159. 10.1016/j.jaac.2012.08.018

Dawson, G., Rogers, S., Munson, J., Smith, M., Winter, J., Greenson, J., Donaldson, A., & Varley, J. (2010). Randomized, controlled trial of an intervention for toddlers with autism: The Early Start Denver Model. Pediatrics, 125(1), e17–23. 10.1542/peds.2009-0958

Eisenhauer, J. G. (2021). Meta-analysis and mega-analysis: A simple introduction. Teaching Statistics, 43(1), 21–27. 10.1111/test.12242

Eldevik, S., Hastings, R. P., Hughes, J. C., Jahr, E., Eikeseth, S., & Cross, S. (2009). Meta-analysis of Early Intensive Behavioral Intervention for children with autism. Journal of Clinical Child and Adolescent Psychology: The Official Journal for the Society of Clinical Child and Adolescent Psychology, American Psychological Association, Division 53, 38(3), 439–450. 10.1080/15374410902851739

Esler, A. N., Bal, V. H., Guthrie, W., Wetherby, A., Ellis Weismer, S., & Lord, C. (2015). The Autism Diagnostic Observation Schedule, Toddler Module: Standardized Severity Scores. Journal of Autism and Developmental Disorders, 45(9), 2704–2720. 10.1007/s10803-015-2432-7

Estes, A., Munson, J., Rogers, S. J., Greenson, J., Winter, J., & Dawson, G. (2015). Long-Term Outcomes of Early Intervention in 6-Year-Old Children With Autism Spectrum Disorder. Journal of the American Academy of Child and Adolescent Psychiatry, 54(7), 580–587. 10.1016/j.jaac.2015.04.005

Fuller, E. A., & Kaiser, A. P. (2020). The Effects of Early Intervention on Social Communication Outcomes for Children with Autism Spectrum Disorder: A Meta-analysis. Journal of Autism and Developmental Disorders, 50(5), 1683–1700. 10.1007/s10803-019-03927-z

Fuller, E. A., Oliver, K., Vejnoska, S. F., & Rogers, S. J. (2020). The Effects of the Early Start Denver Model for Children with Autism Spectrum Disorder: A Meta-Analysis. Brain Sciences, 10(6), 368. 10.3390/brainsci10060368

Gentles, S. J., Ng-Cordell, E. C., Hunsche, M. C., McVey, A. J., Bednar, E. D., DeGroote, M. G., Chen, Y.-J., Duku, E., Kerns, C. M., Banfield, L., Szatmari, P., & Georgiades, S. (2023). Trajectory research in children with an autism diagnosis: A scoping review. Autism: The International Journal of Research and Practice, 13623613231170280. 10.1177/13623613231170280

Godel, M., Robain, F., Kojovic, N., Franchini, M., Wood de Wilde, H., & Schaer, M. (2022). Distinct Patterns of Cognitive Outcome in Young Children With Autism Spectrum Disorder Receiving the Early Start Denver Model. Frontiers in Psychiatry, 13, 835580. 10.3389/fpsyt.2022.835580

Gotham, K., Pickles, A., & Lord, C. (2009). Standardizing ADOS scores for a measure of severity in autism spectrum disorders. Journal of Autism and Developmental Disorders, 39(5), 693–705. 10.1007/s10803-008-0674-3

Griffith, R., Luiz, D., & Association for Research in Infant and Child Development. (2006). Griffiths mental development scales, extended revised: GMDS-ER; two to eight years. Association for Research in Infant and Child Development.

Howlin, P., Magiati, I., & Charman, T. (2009). Systematic review of early intensive behavioral interventions for children with autism. American Journal on Intellectual and Developmental Disabilities, 114(1), 23–41. 10.1352/2009.114:23;nd41

Hus, V., Gotham, K., & Lord, C. (2014). Standardizing ADOS domain scores: Separating severity of social affect and restricted and repetitive behaviors. Journal of Autism and Developmental Disorders, 44(10), 2400–2412. 10.1007/s10803-012-1719-1

Jones, E. J. H., Dawson, G., Kelly, J., Estes, A., & Webb, S. J. (2017). Parent-delivered early intervention in infants at risk for ASD: Effects on electrophysiological and habituation measures of social attention. Autism Research: Official Journal of the International Society for Autism Research, 10(5), 961–972. 10.1002/aur.1754

Lombardo, M. V., Busuoli, E. M., Schreibman, L., Stahmer, A. C., Pramparo, T., Landi, I., Mandelli, V., Bertelsen, N., Barnes, C. C., Gazestani, V., Lopez, L., Bacon, E. C., Courchesne, E., & Pierce, K. (2021). Pre-treatment clinical and gene expression patterns predict developmental change in early intervention in autism. Molecular Psychiatry. 10.1038/s41380-021-01239-2

Lombardo, M. V., Lai, M.-C., & Baron-Cohen, S. (2019). Big data approaches to decomposing heterogeneity across the autism spectrum. Molecular Psychiatry. 10.1038/s41380-018-0321-0

Lombardo, M. V., & Mandelli, V. (2022). Rethinking Our Concepts and Assumptions About Autism. Frontiers in Psychiatry, 13, 903489. 10.3389/fpsyt.2022.903489

Mandelli, V., Landi, I., Busuoli, E. M., Courchesne, E., Pierce, K., & Lombardo, M. V. (2023). Prognostic early snapshot stratification of autism based on adaptive functioning. Nature Mental Health, 1(5), 327–336. 10.1038/s44220-023-00056-6

McGlade, A., Whittingham, K., Barfoot, J., Taylor, L., & Boyd, R. N. (2023). Efficacy of very early interventions on neurodevelopmental outcomes for infants and toddlers at increased likelihood of or diagnosed with autism: A systematic review and meta-analysis. Autism Research: Official Journal of the International Society for Autism Research, 16(6), 1145– 1160. 10.1002/aur.2924

Mullen, E. (1995). Mullen Scales of Early Learning. American Guidance Service.

Muratori, F., Narzisi, A., & IDIA Group. (2014). Exploratory study describing 6 month outcomes for young children with autism who receive treatment as usual in Italy. Neuropsychiatric Disease and Treatment, 10, 577–586. 10.2147/NDT.S58308

Pellicano, E., & den Houting, J. (2021). Annual Research Review: Shifting from “normal science” to neurodiversity in autism science. Journal of Child Psychology and Psychiatry, and Allied Disciplines. 10.1111/jcpp.13534

Pierce, K., Gazestani, V., Bacon, E., Courchesne, E., Cheng, A., Barnes, C. C., Nalabolu, S., Cha, D., Arias, S., Lopez, L., Pham, C., Gaines, K., Gyurjyan, G., Cook-Clark, T., & Karins, K. (2021). Get SET Early to Identify and Treatment Refer Autism Spectrum Disorder at 1 Year and Discover Factors That Influence Early Diagnosis. The Journal of Pediatrics, 236, 179–188. 10.1016/j.jpeds.2021.04.041

Robain, F., Franchini, M., Kojovic, N., Wood de Wilde, H., & Schaer, M. (2020). Predictors of Treatment Outcome in Preschoolers with Autism Spectrum Disorder: An Observational Study in the Greater Geneva Area, Switzerland. Journal of Autism and Developmental Disorders, 50(11), 3815–3830. 10.1007/s10803-020-04430-6

Rodgers, M., Simmonds, M., Marshall, D., Hodgson, R., Stewart, L. A., Rai, D., Wright, K., Ben-Itzchak, E., Eikeseth, S., Eldevik, S., Kovshoff, H., Magiati, I., Osborne, L. A., Reed, P., Vivanti, G., Zachor, D., & Couteur, A. L. (2021). Intensive behavioural interventions based on applied behaviour analysis for young children with autism: An international collaborative individual participant data meta-analysis. Autism: The International Journal of Research and Practice, 25(4), 1137–1153. 10.1177/1362361320985680

Rogers, S. J., Estes, A., Lord, C., Munson, J., Rocha, M., Winter, J., Greenson, J., Colombi, C., Dawson, G., Vismara, L. A., Sugar, C. A., Hellemann, G., Whelan, F., & Talbott, M. (2019). A Multisite Randomized Controlled Two-Phase Trial of the Early Start Denver Model Compared to Treatment as Usual. Journal of the American Academy of Child and Adolescent Psychiatry, 58(9), 853–865. 10.1016/j.jaac.2019.01.004

Rogers, S. J., Yoder, P., Estes, A., Warren, Z., McEachin, J., Munson, J., Rocha, M., Greenson, J., Wallace, L., Gardner, E., Dawson, G., Sugar, C. A., Hellemann, G., & Whelan, F. (2021). A Multisite Randomized Controlled Trial Comparing the Effects of Intervention Intensity and Intervention Style on Outcomes for Young Children With Autism. Journal of the American Academy of Child and Adolescent Psychiatry, 60(6), 710–722. 10.1016/j.jaac.2020.06.013

Sandbank, M., Bottema-Beutel, K., Crowley LaPoint, S., Feldman, J. I., Barrett, D. J., Caldwell, N., Dunham, K., Crank, J., Albarran, S., & Woynaroski, T. (2023). Autism intervention meta-analysis of early childhood studies (Project AIM): Updated systematic review and secondary analysis. BMJ (Clinical Research Ed.), 383, e076733. 10.1136/bmj-2023-076733

Sandbank, M., Bottema-Beutel, K., Crowley, S., Cassidy, M., Dunham, K., Feldman, J. I., Crank, J., Albarran, S. A., Raj, S., Mahbub, P., & Woynaroski, T. G. (2020). Project AIM: Autism intervention meta-analysis for studies of young children. Psychological Bulletin, 146(1), 1–29. 10.1037/bul0000215

Sandbank, M., Bottema-Beutel, K., Crowley, S., Cassidy, M., Feldman, J. I., Canihuante, M., & Woynaroski, T. (2020). Intervention Effects on Language in Children With Autism: A Project AIM Meta-Analysis. Journal of Speech, Language, and Hearing Research: JSLHR, 63(5), 1537–1560. 10.1044/2020_JSLHR-19-00167

Sandbank, M., Pustejovsky, J. E., Bottema-Beutel, K., Caldwell, N., Feldman, J. I., Crowley LaPoint, S., & Woynaroski, T. (2024). Determining Associations Between Intervention Amount and Outcomes for Young Autistic Children: A Meta-Analysis. JAMA Pediatrics, 178(8), 763–773. 10.1001/jamapediatrics.2024.1832

Sinai-Gavrilov, Y., Gev, T., Mor-Snir, I., Vivanti, G., & Golan, O. (2020). Integrating the Early Start Denver Model into Israeli community autism spectrum disorder preschools: Effectiveness and treatment response predictors. Autism: The International Journal of Research and Practice, 24(8), 2081–2093. 10.1177/1362361320934221

Smith, C. J., James, S., Skepnek, E., Leuthe, E., Outhier, L. E., Avelar, D., Barnes, C. C., Bacon, E., & Pierce, K. (2022). Implementing the Get SET Early Model in a Community Setting to Lower the Age of ASD Diagnosis. Journal of Developmental and Behavioral Pediatrics: JDBP, 43(9), 494–502. 10.1097/DBP.0000000000001130

Smith, T., Klorman, R., & Mruzek, D. W. (2015). Predicting Outcome of Community-Based Early Intensive Behavioral Intervention for Children with Autism. Journal of Abnormal Child Psychology, 43(7), 1271–1282. 10.1007/s10802-015-0002-2

Sparrow, S., Balla, D., Cicchetti, D. V., & Doll, E. A. (2005). Vineland-II Scales Of Adaptive Behavior. American Guidance Service.

Tager-Flusberg, H., & Kasari, C. (2013). Minimally verbal school-aged children with autism spectrum disorder: The neglected end of the spectrum. Autism Research: Official Journal of the International Society for Autism Research, 6(6), 468–478. 10.1002/aur.1329

Vivanti, G., Dissanayake, C., Duncan, E., Feary, J., Capes, K., Upson, S., Bent, C. A., Rogers, S. J., Hudry, K., & Victorian ASELCC Team. (2019). Outcomes of children receiving Group-Early Start Denver Model in an inclusive versus autism-specific setting: A pilot randomized controlled trial. Autism: The International Journal of Research and Practice, 23(5), 1165–1175. 10.1177/1362361318801341

Vivanti, G., Paynter, J., Duncan, E., Fothergill, H., Dissanayake, C., Rogers, S. J., & Victorian ASELCC Team. (2014). Effectiveness and feasibility of the early start denver model implemented in a group-based community childcare setting. Journal of Autism and Developmental Disorders, 44(12), 3140–3153. 10.1007/s10803-014-2168-9

Vivanti, G., Prior, M., Williams, K., & Dissanayake, C. (2014). Predictors of outcomes in autism early intervention: Why don’t we know more? Frontiers in Pediatrics, 2, 58. 10.3389/fped.2014.00058

Waizbard-Bartov, E., Ferrer, E., Young, G. S., Heath, B., Rogers, S., Wu Nordahl, C., Solomon, M., & Amaral, D. G. (2021). Trajectories of Autism Symptom Severity Change During Early Childhood. Journal of Autism and Developmental Disorders, 51(1), 227–242. 10.1007/s10803-020-04526-z

Warren, Z., McPheeters, M. L., Sathe, N., Foss-Feig, J. H., Glasser, A., & Veenstra-Vanderweele, J. (2011). A systematic review of early intensive intervention for autism spectrum disorders. Pediatrics, 127(5), e1303–1311. 10.1542/peds.2011-0426

Zwaigenbaum, L., Bauman, M. L., Choueiri, R., Kasari, C., Carter, A., Granpeesheh, D., Mailloux, Z., Smith Roley, S., Wagner, S., Fein, D., Pierce, K., Buie, T., Davis, P. A., Newschaffer, C., Robins, D., Wetherby, A., Stone, W. L., Yirmiya, N., Estes, A., … Natowicz, M. R. (2015). Early Intervention for Children With Autism Spectrum Disorder Under 3 Years of Age: Recommendations for Practice and Research. Pediatrics, 136 Suppl 1(Suppl 1), S60–81. 10.1542/peds.2014-3667E

